# Post-operative serum procalcitonin vs C reactive Protein as a marker of post-operative infectious complications in pancreatic surgery – A systemic review and meta-analysis

**DOI:** 10.1101/2020.10.06.20208181

**Authors:** Bhavin Vasavada, Hardik Patel

## Abstract

**Aim of Study:** Aim of this meta-analysis was to compare diagnostic accuracy of C reactive Protein and Procalcitonin between postoperative day 3 to 5 in predicting infectious complications post pancreatic surgery.

**Methods:** Systemic literature search was performed using MEDLINE, EMBASE and SCOPUS to identify studies evaluating the diagnostic accuracy of Procalcitonin (PCT) and C-Reactive Protein (CRP) as a predictor for detecting infectious complications between postoperative days (POD) 3 to 5 following pancreatic surgery. A meta-analysis was performed using random effect model and pooled predictive parameters. Geometric means were calculated for PCT cut offs. The work has been reported in line with PRISMA guidelines.

**Results:** After applying inclusion and exclusion criteria 15 studies consisting of 2212 patients were included in the final analysis according to PRISMA guidelines. Pooled sensitivity, specificity, Area under curve and diagnostic odds ratio (DOR)for day 3 C-reactive protein was respectively 62%,67% 0.772 and 6.54. Pooled sensitivity, specificity, Area under curve and diagnostic odds ratio (DOR)for day 3 procalcitonin was respectively 74%,79%,0.8453 and 11.03. Sensitivity, specificity, Area under curve, and Diagnostic odds ratio for day 4 C-reactive protein was respectively 60%,68%, 0.8022 and 11.90. Pooled Sensitivity, specificity and Diagnostic odds ratio of post-operative day 5 procalcitonin level in predicting infectious complications were respectively 83%,70% and 12.9. Pooled Sensitivity, specificity, AUROC and diagnostic odds ratio were respectively 50%,70%, 0.777 and 10.19.

**Conclusion:** Post-operative procalcitonin is better marker to predict post-operative infectious complications after pancreatic surgeries and post-operative day 3 procalcitonin has highest diagnostic accuracy.

## Introduction

Pancreatic surgeries (Pancreaticoduodenectomy/ distal pancreatectomy) are the main treatments for various benign and malignant disease of pancreas, duodenum, and ampullary region. [1]. Pancreatic surgeries are still associated with very high morbidity and mortality. [2]. Majority of complications following pancreatic surgeries are infectious complications including pancreatic leaks and fistula. [3]. These complications can affect outcomes and also increase cost for pancreatic surgeries. [4].

C reactive protein (CRP) and procalcitonin are suggested as inflammatory markers for diagnosing infective complications following colorectal and abdominal surgeries. [5-10]. CRP is not considered as a specific marker for infection, as it can rise in any inflammatory condition. [11].

Procalcitonin is now emerging as a useful and specific marker for sepsis and guide to antibiotic treatment. [12]. It is suggested as a useful marker in predicting infectious complications for colorectal surgeries. [5].

However, there is still limited literature comparing effectiveness of C-Reactive Protein and Procalcitonin (PCT) as a marker of infectious complications post pancreatic surgeries and very few studies to show which is better marker to diagnose infectious complications.

Pancreatic surgeries are highly morbid surgeries where early diagnosis of complications can help to reduce mortality.

## AIM of the study

Aim of this meta-analysis was to compare diagnostic accuracy of C reactive Protein and Procalcitonin between postoperative day 3 to 5 in predicting infectious complications post pancreatic surgery.

## Materials and Methods

### Data collection

Medline (PubMed), Embase and Scopus were searched with key words like “procalcitonin”, “C reactive Protein”, “pancreatic surgery”, “pancreaticoduodenectomy”, “distal pancreatectomy”, “post-operative complications”, “infective complication”, “pancreatic leak”, “pancreatic fistula”, “anastomotic leak”. Studies after Year 2005 (last 15 years) were searched. Anastomotic leak and pancreatic fistula were considered as infectious complications and were included in search strategy. The work has been reported in line with PRISMA (Preferred Reporting Items for Systemic Reviews) and MOOSE (Meta-analysis of observational studies in epidemiology) guidelines. [13,14]

### Definition of post-operative infectious complications

Infectious complications were defined as any complications like intraabdominal abscess, pancreatic leak, pancreatic fistula, wound complications, urinary tract infection, post-operative pneumonia or adult respiratory distress syndrome. Only clinically significant pancreatic fistula (ISGPS grade b/c) was considered as an infectious complication. [15] Screening was done by two reviewers (BV and HP) independently at the title, abstract, and full text stages. Any disagreements were discussed between the reviewers before a final decision was made.

### Study selection

#### Inclusion criteria

- Randomized control trials
- Observational cohort study
- Studies which included post-operative procalcitonin or C-reactive protein level between postoperative day 3 to 5.
- Studies where subject underwent pancreaticoduodenectomy or distal pancreatectomy
- Studies which included patients with age 18 and above.
- Studies which evaluated post-operative complications.

#### Exclusion criteria

- Studies where full text articles could not be obtained.
- Studies which included only post-operative day 1,2 or pre-operative procalcitonin or C-reactive protein level.

#### Data extraction

Information on study characteristics including patient population, study duration, follow-up period, index test, and reference standard were extracted from each study. The primary outcome, i.e., diagnostic performance of PCT or CRP to detect infectious complications reported as sensitivity (Se), specificity (Sp), positive predictive value (PPV), negative predictive value (NPV), positive likelihood ratio (LR+), and negative likelihood ratio (LR−) at POD 3 and 5, was collected. As anastomotic leakage or pancreatic fistula were considered a subset of infectious complications and expected to account for most cases of infectious complications in pancreatic surgery, it was used as the surrogate outcome of interest during data extraction in studies which did not specifically report infectious complications.

Raw data from the articles were used to construct 2*2 tables (true positive, false positive, true negative, and false negative). When unavailable, the tables were constructed using the sensitivity and specificity values provided. For each study, the sensitivity and specificity values mentioned in the article were verified by the reconstruction of the 2*2 contingency table using the data specified in the article.

#### Risk of bias assessment

The revised Quality Assessment of Diagnostic Accuracy Studies (QUADAS-2) tool developed by the Cochrane Collaboration was used to assess for the risk of bias and applicability of each study. [16].

The tool consists of four key domains, i.e., patient selection, index test, reference standard, and patient flow through the study and timing of tests. Two reviewers (BV and HP) assessed the study quality independently. In case of disagreement, the judgment was discussed among themselves before a final decision. publication bias was assessed with the Deeks test. [17].

#### Statistical analysis

The statistical analysis was performed according to the Preferred Report Items for Systematic Reviews and Meta-analysis (PRISMA) statement. [13]. The pooled prevalence of infectious complications with corresponding 95% confidence interval (95% CI) was calculated using random effect model. The pooled PCT and CRP cut-off value was derived using geometric mean of the reported PCT and CRP cut-off values. [17]. Using a random effect model, the pooled Se, Sp, LR+, LR−, and diagnostic odds ratios (DOR) with corresponding 95% CI were calculated. Symmetrical summary receiver operating characteristic (SROC) curves were also generated. The area under the curve (AUC) and Q* index (the point on the SROC curve where Se and Sp were equal) were calculated, respectively.[18]. Heterogeneity was assessed using the Higgins I2 test, with values of 25, 50, and 75% indicating low, moderate, and high degrees of heterogeneity, respectively. [19]. Meta-regression and subgroup analyses were attempted whenever feasible.

The statistical analysis was performed using Meta-DiSc 1.4 (Hospital Ramon y Cajal and Universidad Complutense de Madrid, Madrid, Spain) and revman 5.4.

## RESULTS

### Data extraction, Study characteristics and quality assessment

“PUBMED”, “SCOPUS”, “EMBASE” database were searched using key words and search strategy described above. Initially 537 studies were screened. After exclusion of duplicates and unrelated studies 86 studies were thoroughly screened. After applying inclusion and exclusion criteria 15 studies consisting of 2212 patients were included in the final analysis according to PRISMA guidelines. [Figure 1]. [10,20-33]

**Figure 1:**
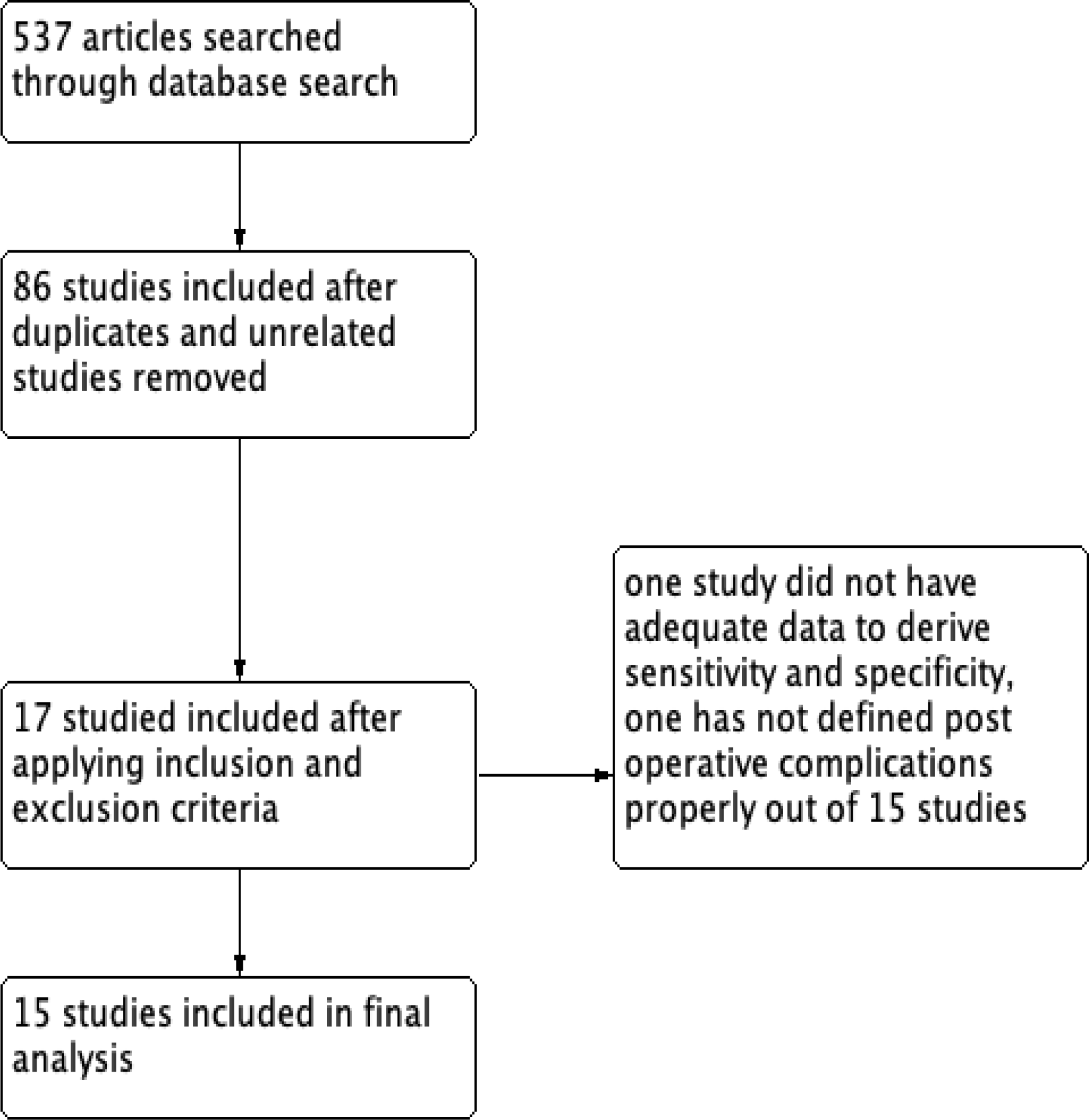
Prisma Flow diagram.

6 studies included analysis of Post-operative day 3 procalcitonin analysis [20-25], 8 studies day 3 C-reactive Protein analysis. 5 studies Included analysis of CRP of day 4. 3 studies included day 3 CRP analysis and 2 studies included day 5 procalcitonin analysis.

Study containing procalcitonin analysis included 471 patients and study containing CRP included 1965 patients. The main characteristics of the included studies are summarized in Table 1. The results of the quality assessment using the QUADAS-2 are shown in Figure. 2. Flaw and timings were unclear in majority of studies.

**Table 1:**
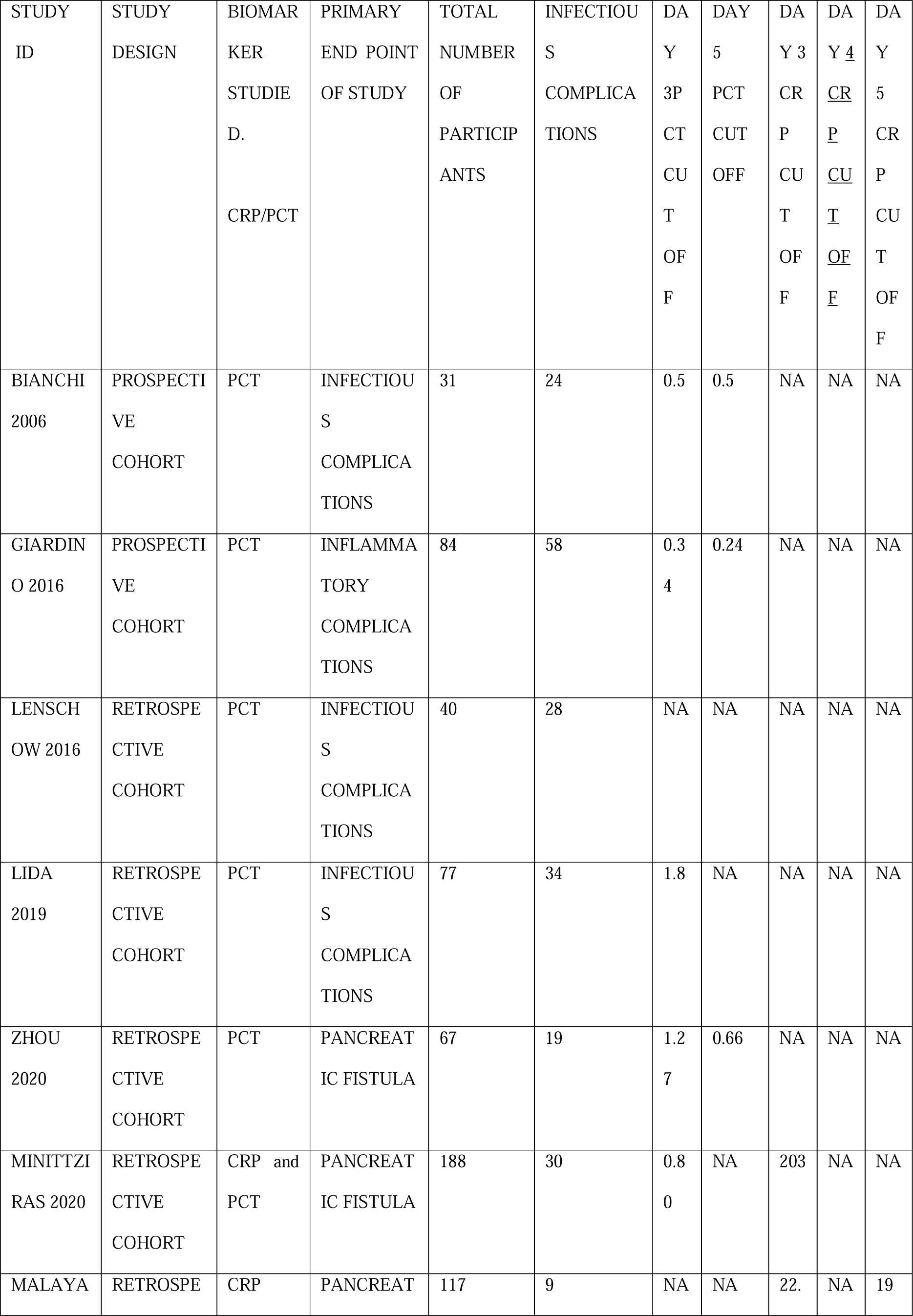
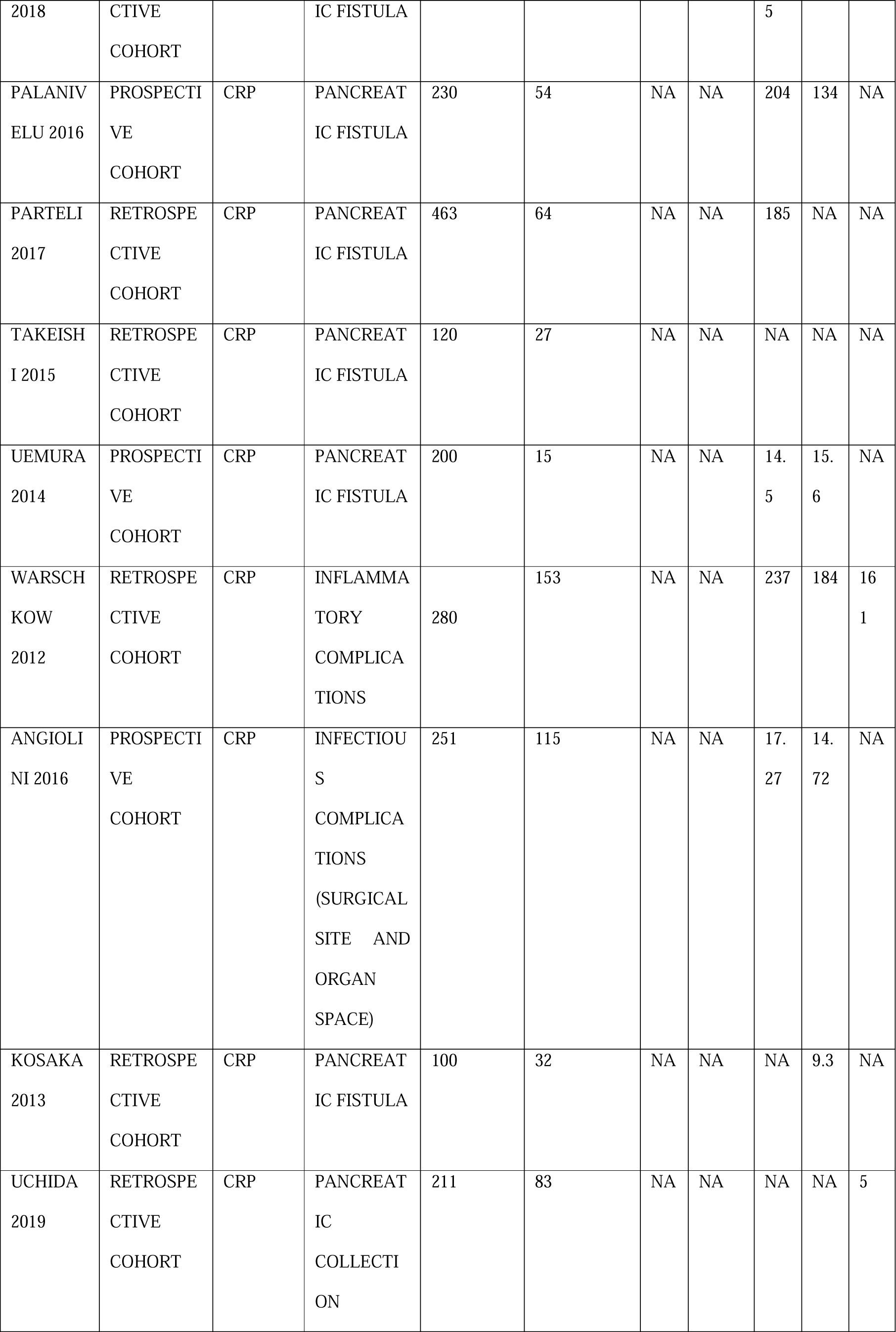
Study characteristics

**Figure 2:**
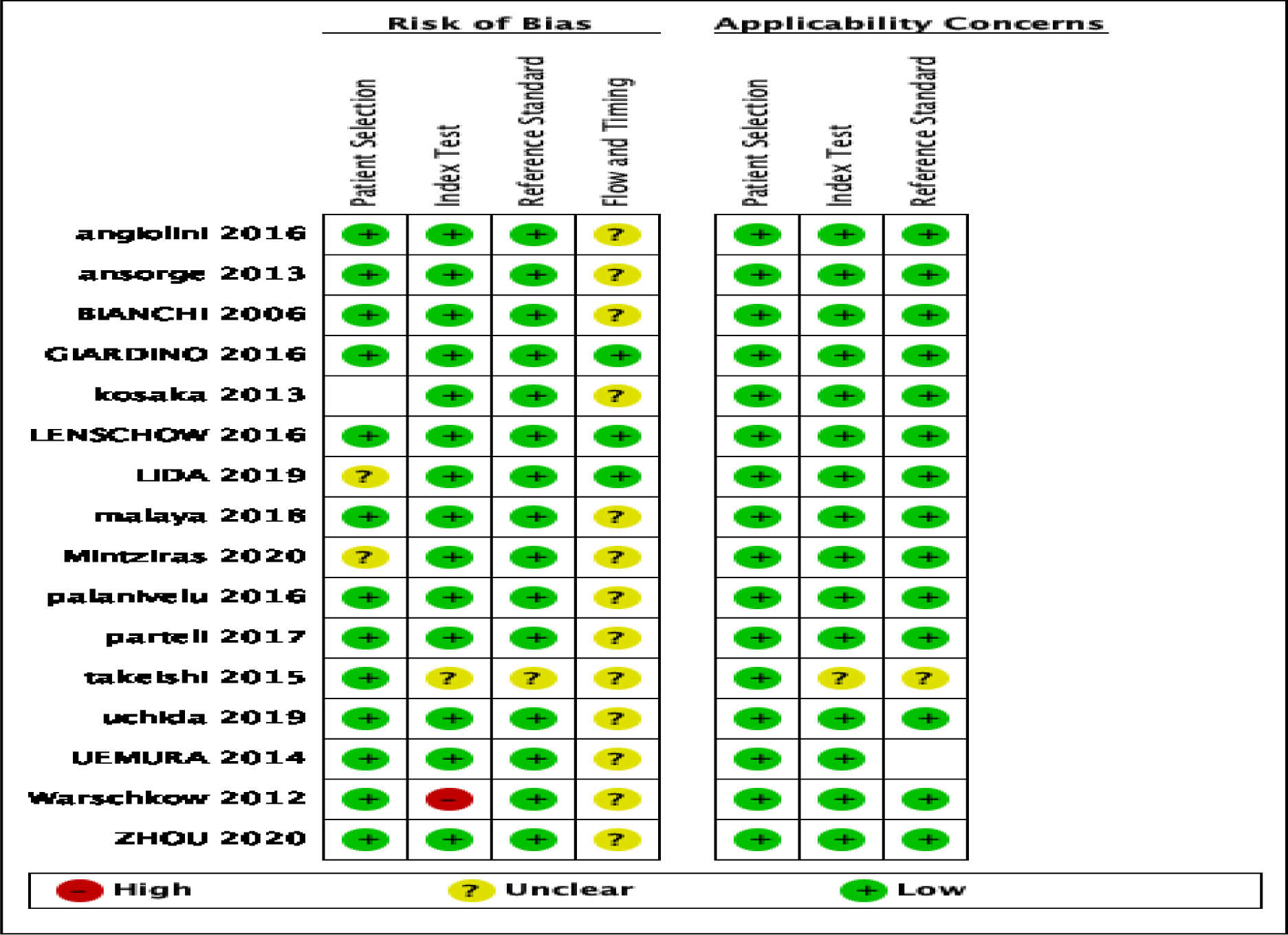
Risk of bias and applicability concerns summary: review authors’ judgements about each domain for each included study

## DIAGNOSTIC ACCURACY ANALYSIS OF POST OPERATIVE DAY 3 C-REACTIVE PROTEIN AND PROCALCITONIN IN PREDICTING INFECTIOUS COMPLICATIONS POST PANCREATIC SURGERY. [FIGURE 3]

Six studies consisting of 465 patients evaluated post-operative day 3 procalcitonin as a marker of infectious complications and 8 studies consisting of 1745 patients evaluated role of post-operative day 3 C-reactive protein as a marker of post-operative infectious complications.

**Figure 3 (a).**
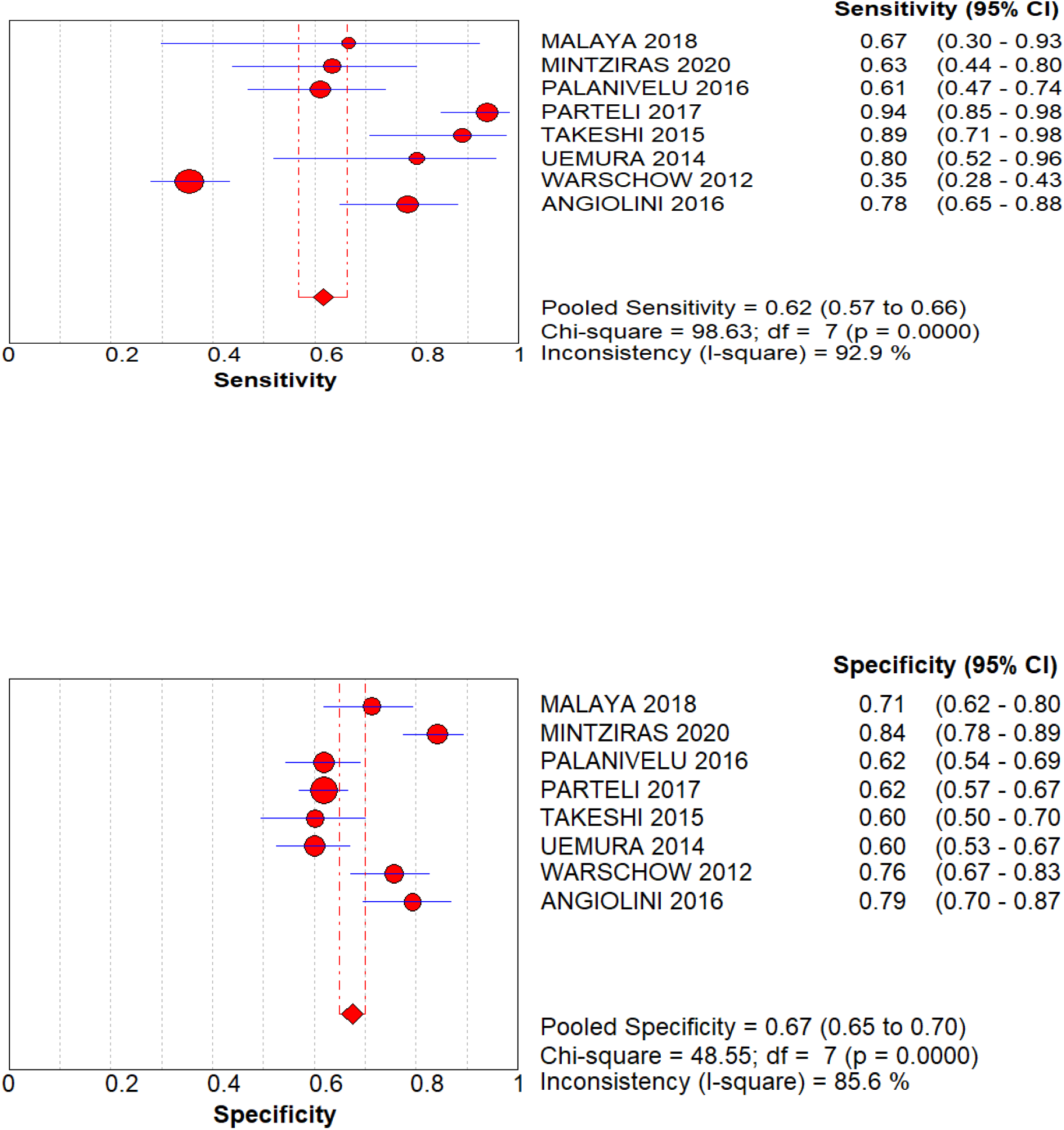
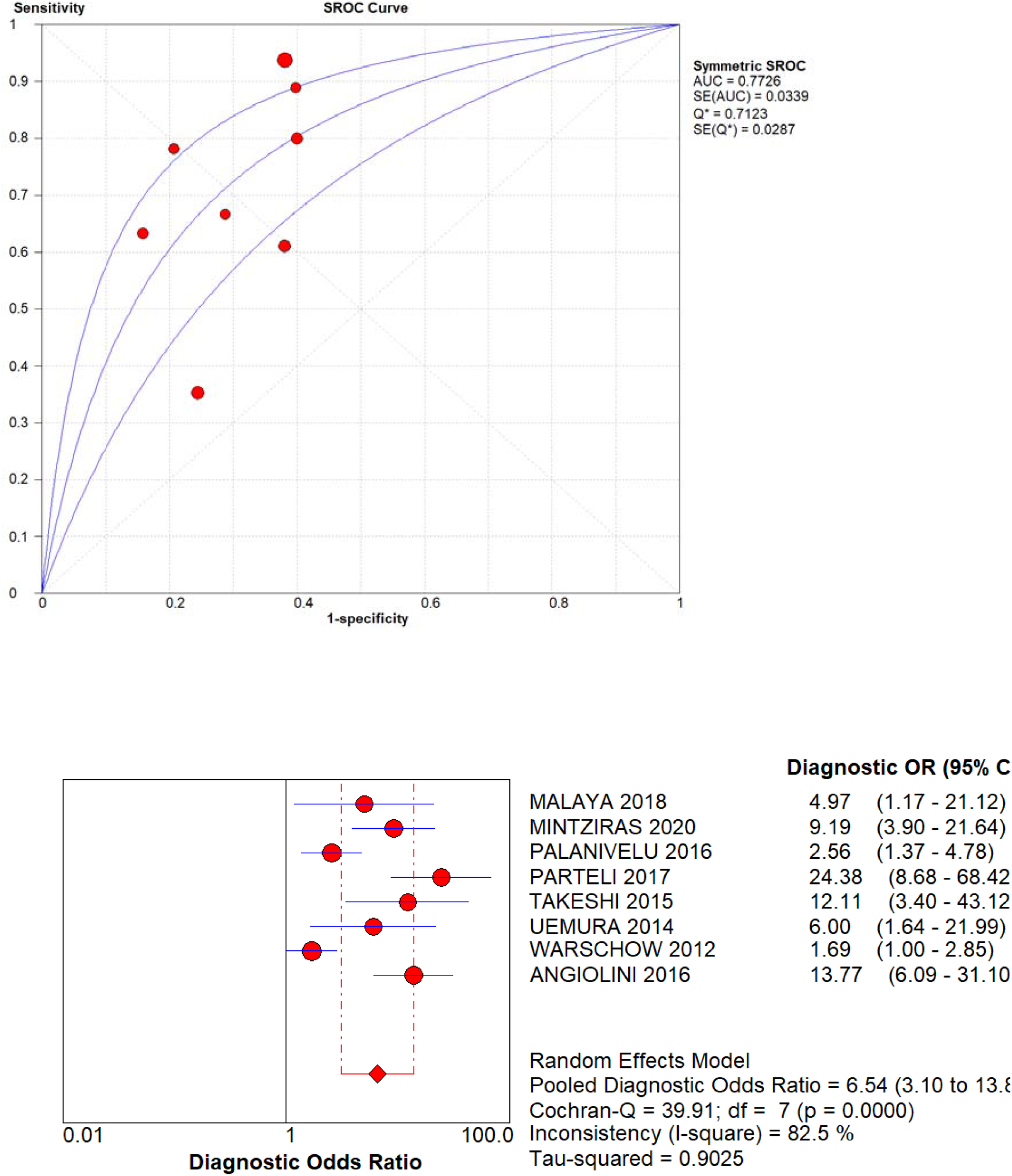
sensitivity, specificity and SROC curve, Diagnostic odds ratio of day 3 CRP as a predictor

Pooled sensitivity, specificity, Area under curve and diagnostic odds ratio (DOR)for day 3 C-reactive protein was respectively 62%,67% 0.772 and 6.54. [Figure 3(a)].

Pooled sensitivity, specificity, Area under curve and diagnostic odds ratio (DOR)for day 3 procalcitonin was respectively 74%,79%,0.8453 and 11.03.[figure 3(b)].

**Figure 3 (b).**
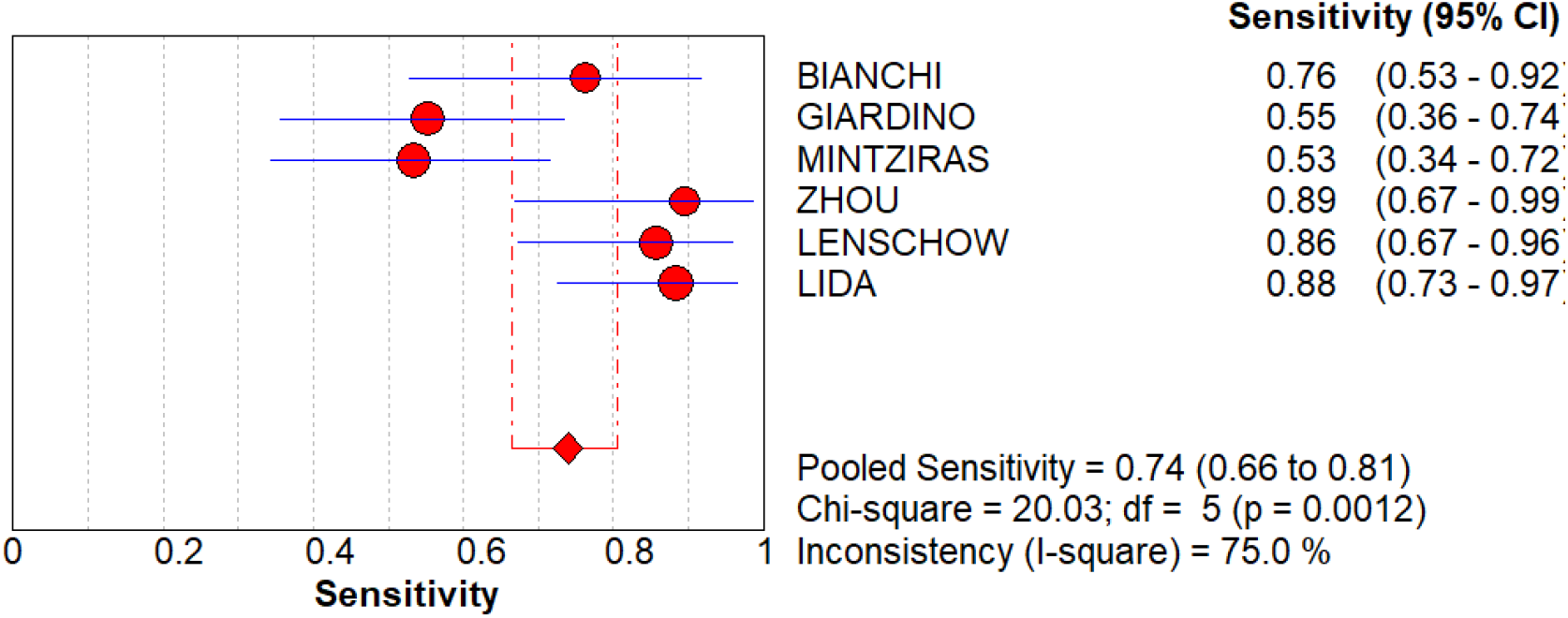
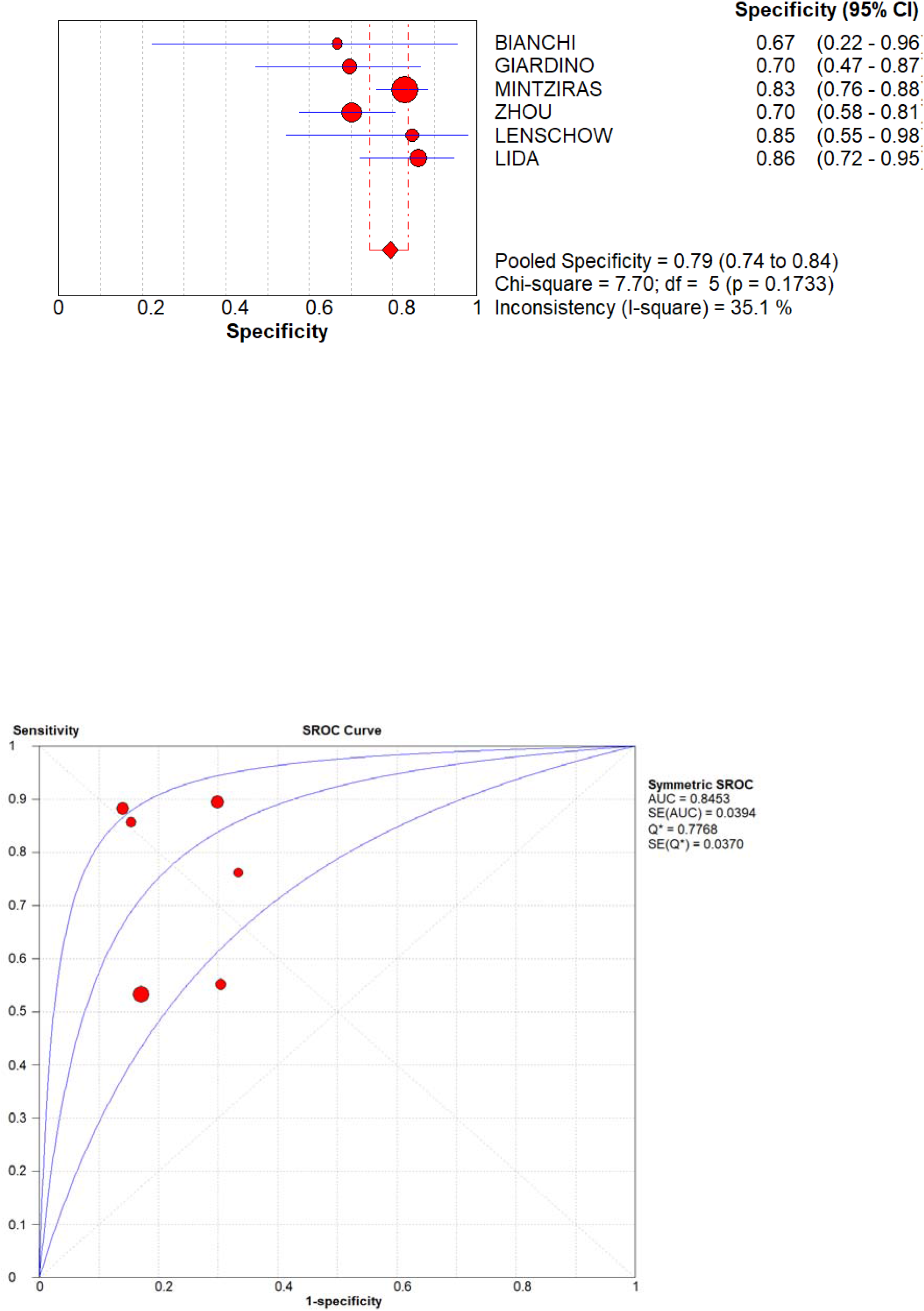
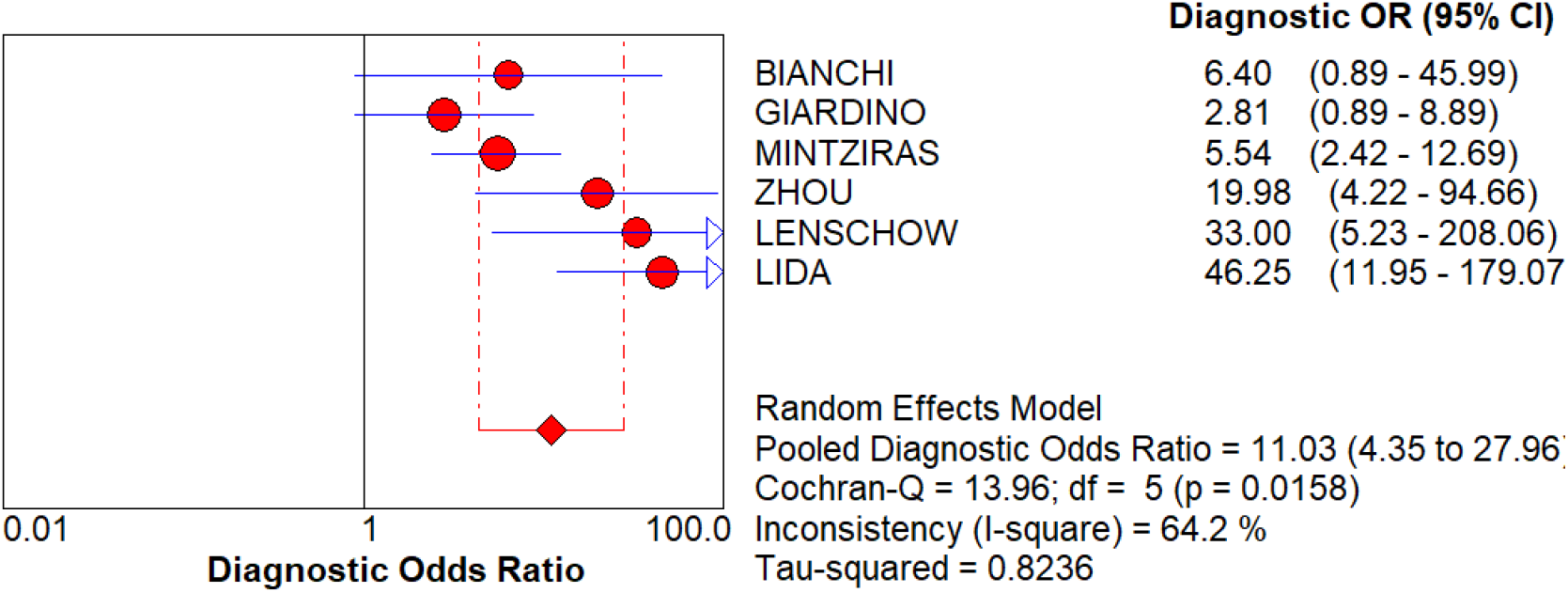
Sensitivity, specificity, SROC curve and Diagnostic ODDS ratio of day 3 Procalcitonin.

## DIAGNOSTIC ACCURACY ANALYSIS OF POST OPERATIVE DAY 4 C-REACTIVE PROTEIN [FIGURE 4]

Five studies consisting of 907 patients evaluated postoperative day 4 C-reactive protein as marker of infectious complications. Sensitivity, specificity, Area under curve, and Diagnostic odds ratio for day 4 C-reactive protein was respectively 60%,68%, 0.8022 and 11.90. No studies evaluated day 4 PCT levels.

**FIGURE. 4:**
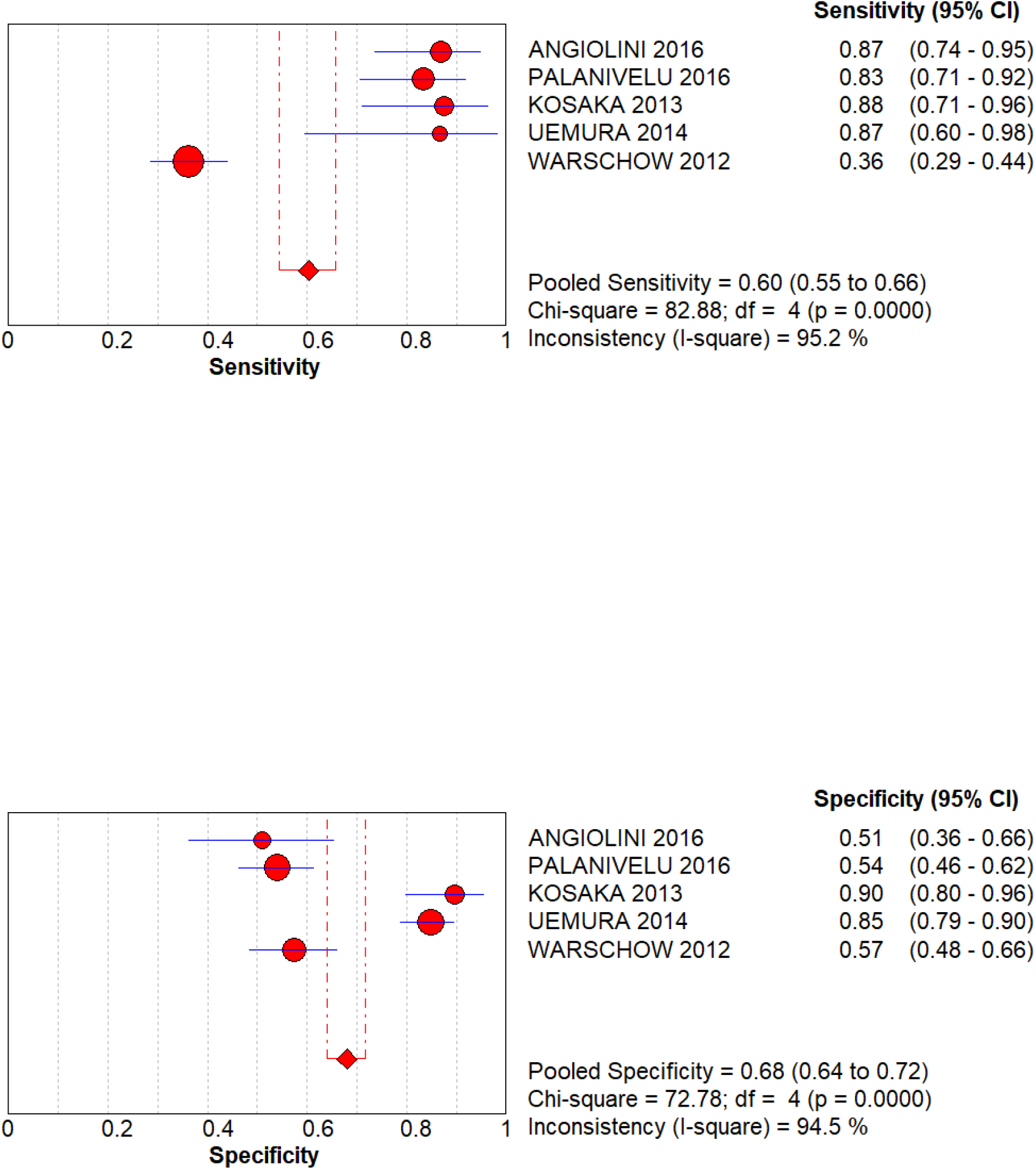
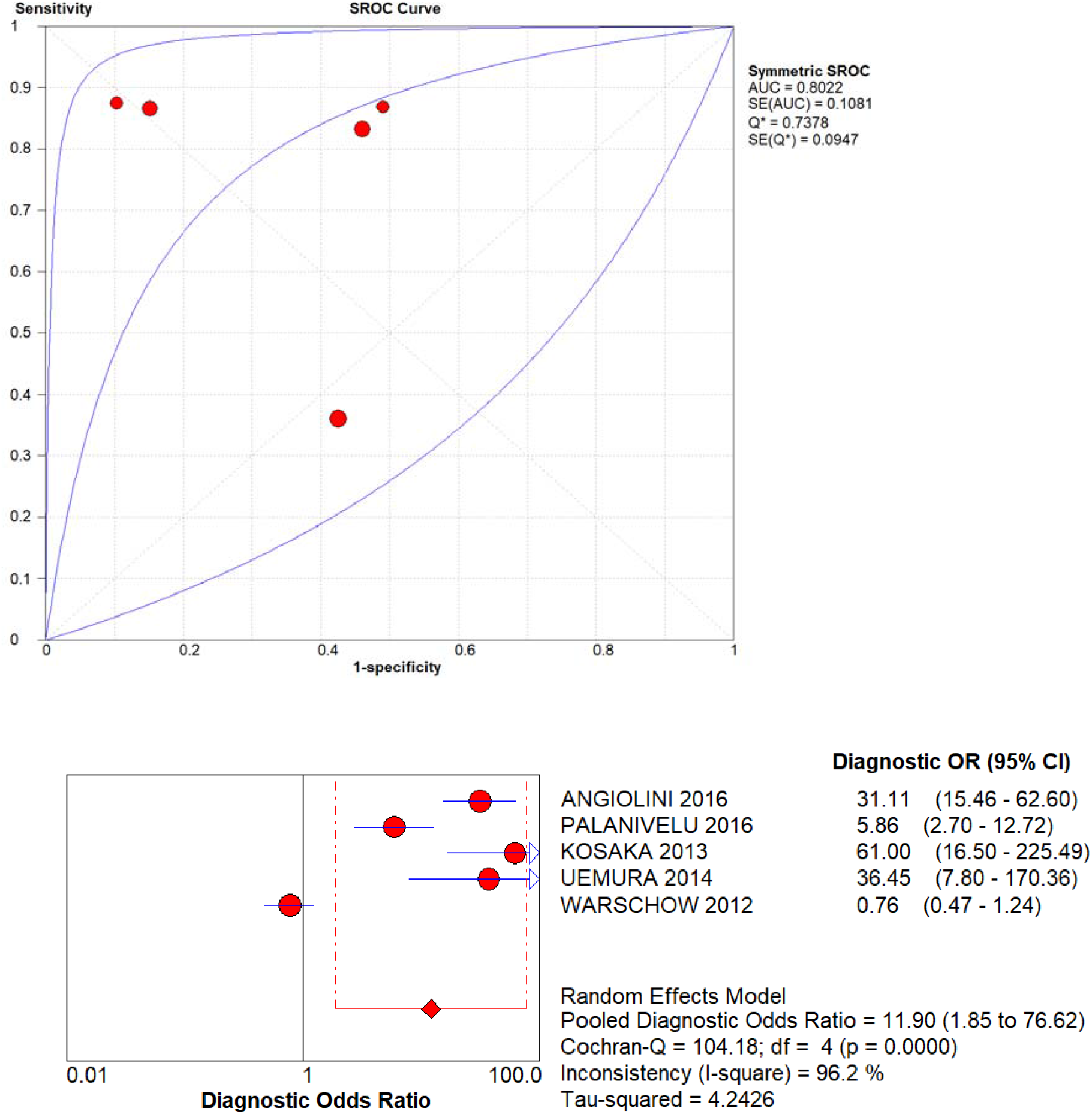
sensitivity, specificity, SROC and DOR of day 4 C-reactive protein.

## DIAGNOSTIC ACCURACY ANALYSIS OF POST OPERATIVE DAY 5 C-REACTIVE PROTEIN AND PROCALCITONIN IN PREDICTING INFECTIOUS COMPLICATIONS POST PANCREATIC SURGERY. [FIGURE 5]

Two studies consisting of 111 patients evaluated post-operative day 5 procalcitonin levels. Pooled Sensitivity, specificity and Diagnostic odds ratio of post-operative day 5 procalcitonin level in predicting infectious complications were respectively 83%,70% and 12.9. SROC could not be constructed as only 2 studies mentioned day 5 procalcitonin levels.

**Figure 5 (a).**
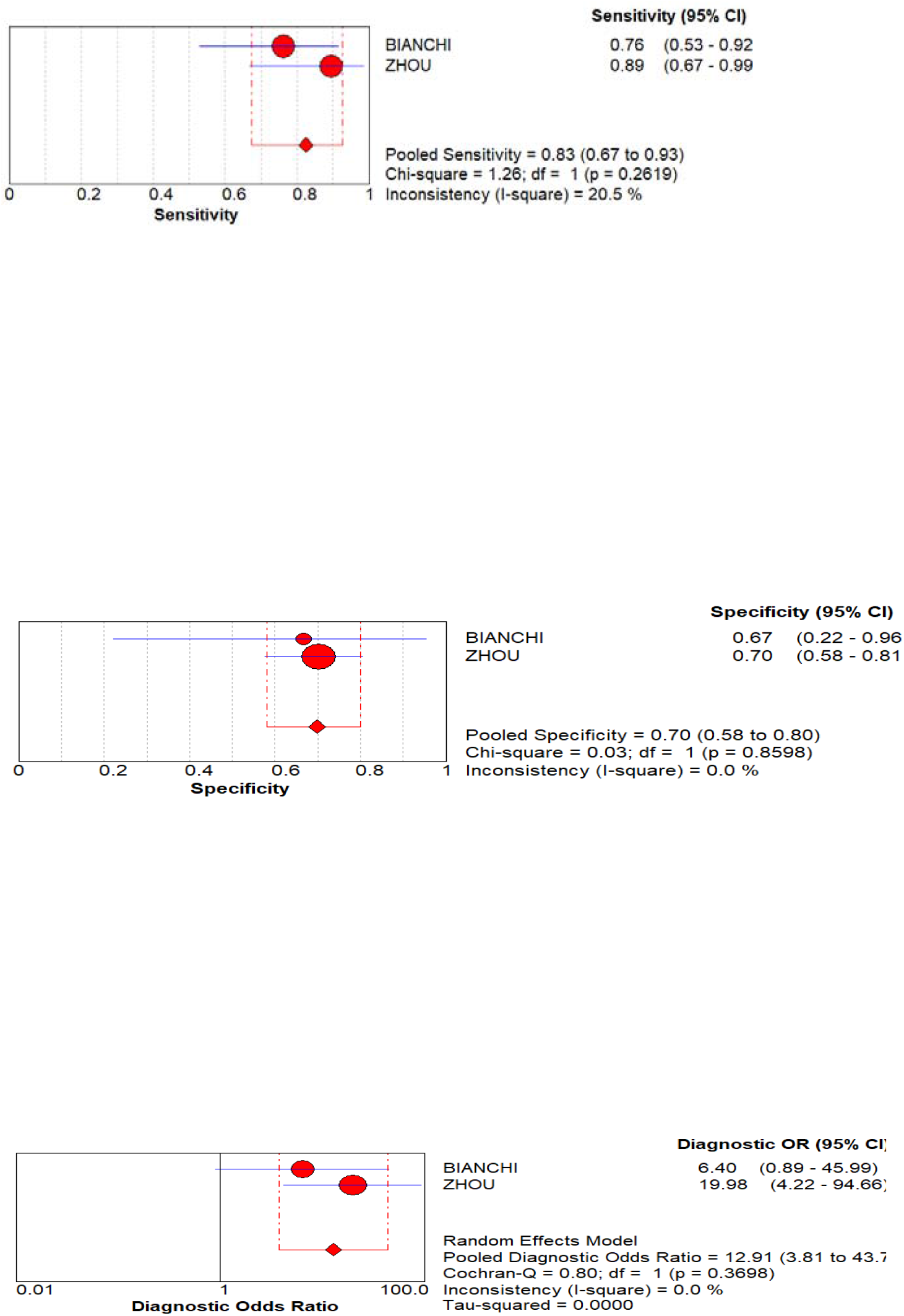
Sensitivity, Specificity and Diagnostic Odds ratio of post-operative day 5 Procalcitonin.

**Figure 5 (b).**
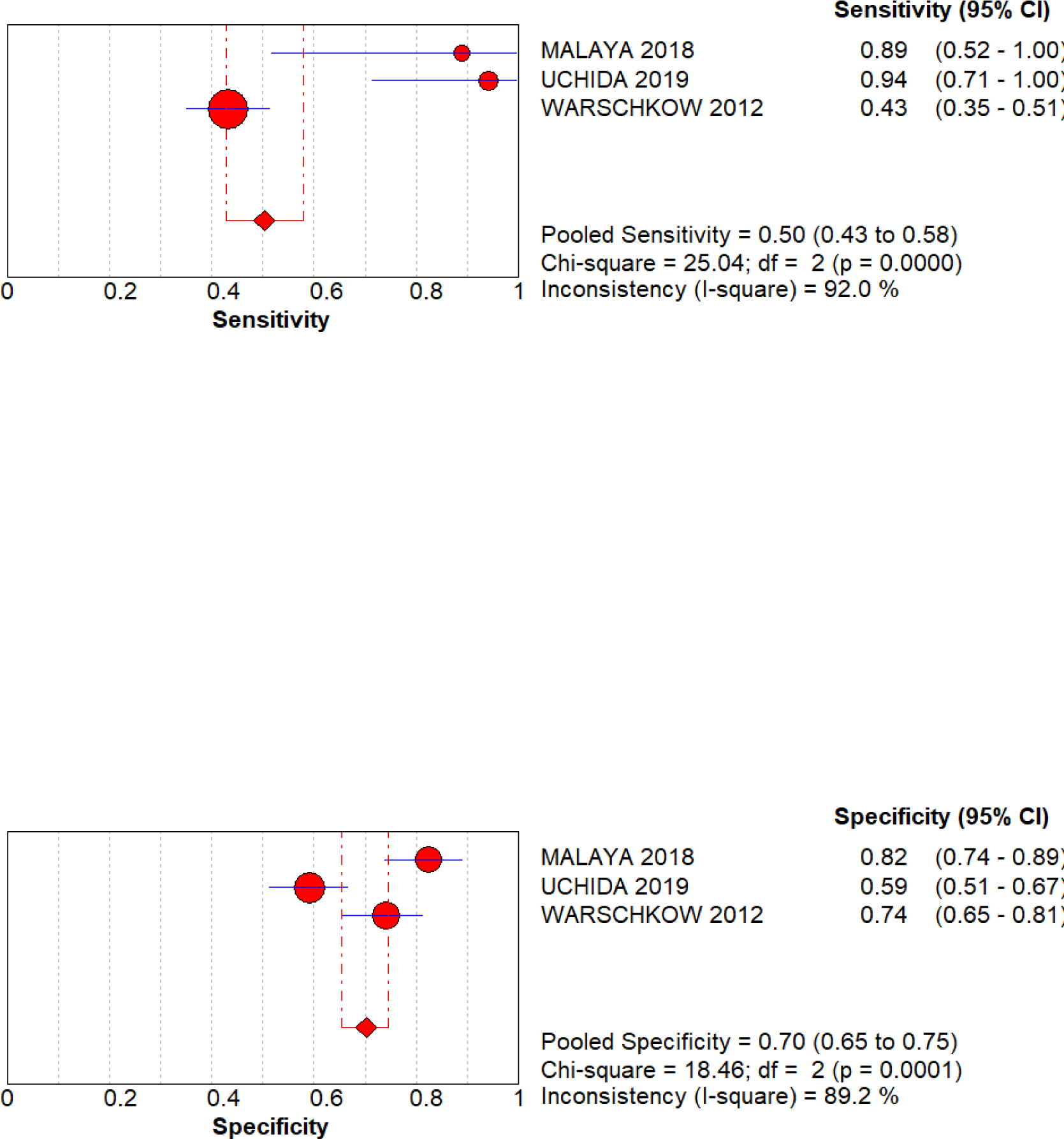
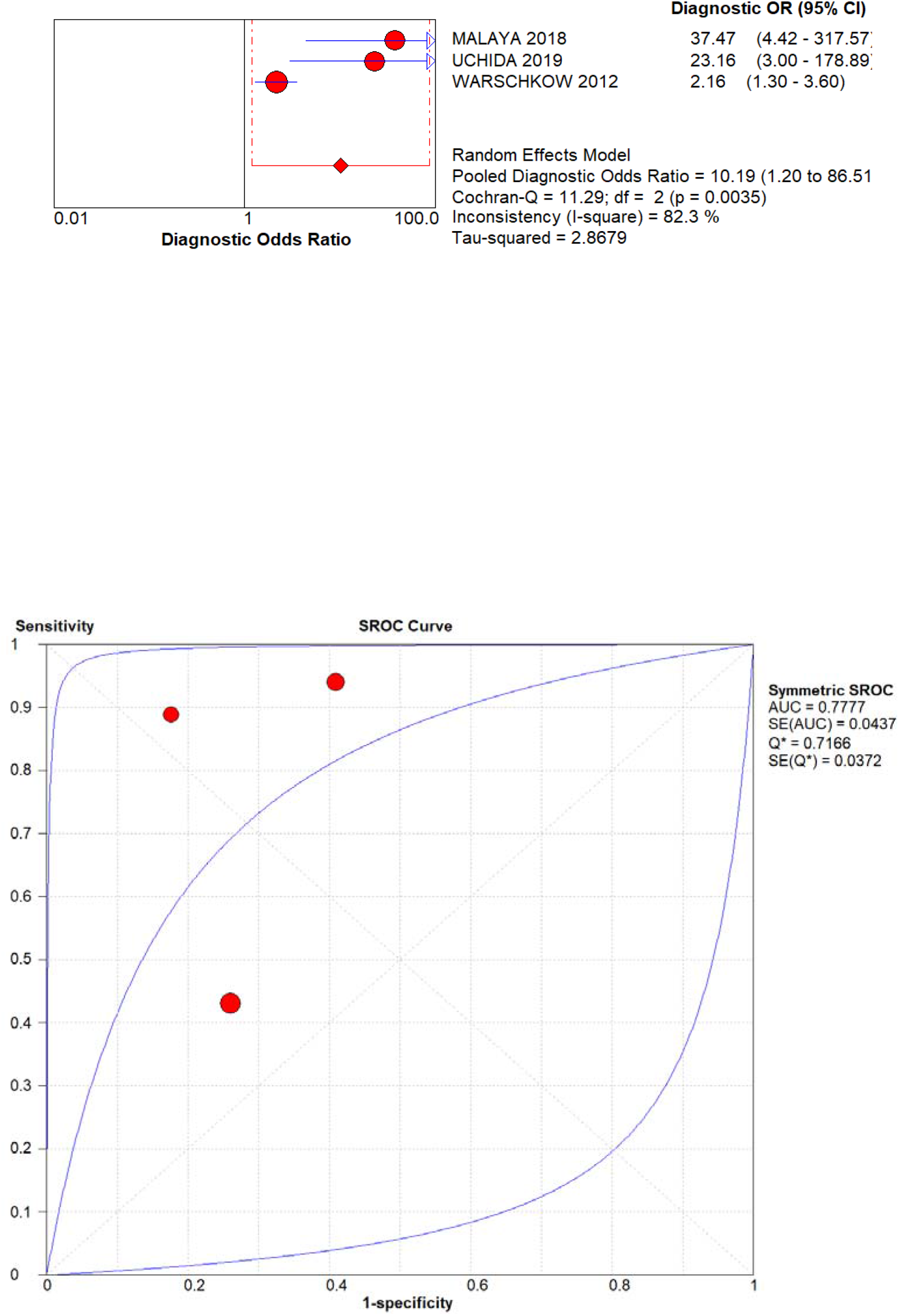
Sensitivity, Specificity, DOR and SROC of post-operative day 5 C reactive protein.

3 studies consisting of 578 patients evaluated post-operative day 5 C-reactive protein as a diagnostic marker for infectious complications after pancreatic surgery. Pooled Sensitivity, specificity, AUROC and diagnostic odds ratio were respectively 50%,70%, 0.777 and 10.19.

## POSITIVE AND NEGATIVE LIKE HOOD RATIO. [SUPPLEMENT FIGURE 1 AND 2]

Pooled positive like hood ratios for post-operative day 3,4 and 5 C-reactive protein were respectively 2.29,2.53,2.62. Pooled Negative like hood ratios of day 3,4,5 CRP were 0.37,0.27,0.25.

Pooled positive like hood ratios for post-operative day 3 and 5 procalcitonin were respectively 3.17 and 2.91. Pooled Negative like hood ratios of day 3 and. 5 Procalcitonin were 0.31 and 0.25.

### C-reactive protein and Procalcitonin cut off

Geometric mean PCT cut off for predicting infectious complications at day 3 was 0.80 with 95% C.I. 0.58-1.02. Geometric mean PCT cut off for predicting infectious complications at day 5 was 0.43 with 95% C.I. 0.20-0.65.

Geometric mean CRP cut off for predicting infectious complications at day 3 was 72.2 with 95% C.I. 2-142. Geometric mean CRP cut off for predicting infectious complications at day 4 was 25.3 with 95% C.I. 0-97. Geometric mean CRP cut off for predicting infectious complications at day 5 was 24.8 with 95% C.I. 0-104.

Deek test for publication bias was not significant. (p=0.456)

## DISCUSSION

In our meta-analysis we evaluated role of Post-operative C-reactive protein and Procalcitonin in predicting post-operative infectious complications. Tan et al. [5] and cousin et al. [34] had done similar meta-analysis showing use of PCT as a predictor for infectious complications following colorectal surgeries. However, to our knowledge this is the first diagnostic accuracy meta-analysis which simultaneously analysed role of .C-reactive protein (CRP) and procalcitonin (PCT).

Survival Sepsis Guidelines 2016.[35] suggests use of PCT as a marker for diagnosing sepsis as well as marker for de-escalation of antibiotics and its use in management of sepsis is gaining popularity now. We decided to use PCT levels at day 3 and day 5 as evidences suggests that PCT can be falsely elevated in first 2 post-operative days. [36,37,38]. We found no study that reported day 4 PCT.

CRP is a known inflammatory marker, however CRP levels can rise in multiple inflammatory condition. We here evaluated day 3,4,5 CRP levels for the same reason as in initial post-operative days surgical stress itself can cause elevated CRP levels.

Highest pooled sensitivity, Diagnostic odds ratio, pooled area under curve for CRP in detecting infectious complications were highest on 4^th^ post-operative day which was respectively 60%, 11.90 and 0.8022. Highest pooled specificity was on 5^th^ post-operative day, which was 70%.

For procalcitonin pooled sensitivity, specificity, pooled area under curve was on post-operative day 3 which were respectively 74%,79%,0.8453 and 11.03. Pooled sensitivity, specificity and diagnostic odds ratios for day 5 procalcitonin were 83%,70% and 12.9. However only 2 studies evaluated post-operative day 5 procalcitonin levels so pooled area under curve could not be calculated. From above findings it seems that post-operative procalcitonin is more sensitive and specific than C-reactive protein in predicting post-operative infectious complications after pancreatic surgeries. Post-operative day 3 procalcitonin is found to be more accurate marker of post-operative infectious complications after pancreatic surgery.

There were certain limitations of these analysis, first is that end point was not similar in every study. Some study evaluated infectious complications and majority evaluated pancreatic leak and fistula. We considered pancreatic fistula as an infectious complication. Heterogenicity was moderate to high in some analysis. Day 5 analysis included very small number of studies. Another limitation is majority of studies included pancreaticoduodenectomies only so to confirm these findings in distal pancreatectomies including laparoscopic distal pancreatectomies we need more data.

However, to best of our knowledge this is the only meta-analysis in which an humble attempt is done to compare CRP and PCT as predictive markers for post0operative infectious complications after pancreatic surgeries.

In conclusion, it shows post-operative procalcitonin is better marker to predict post-operative infectious complications after pancreatic surgeries and post-operative day 3 procalcitonin has highest diagnostic accuracy.

## Data Availability

data will be provided on demand

## Abbreviations

(CRP): C-Reactive Protein
(PCT): Procalcitonin

---

**Supplement Figure 1:**
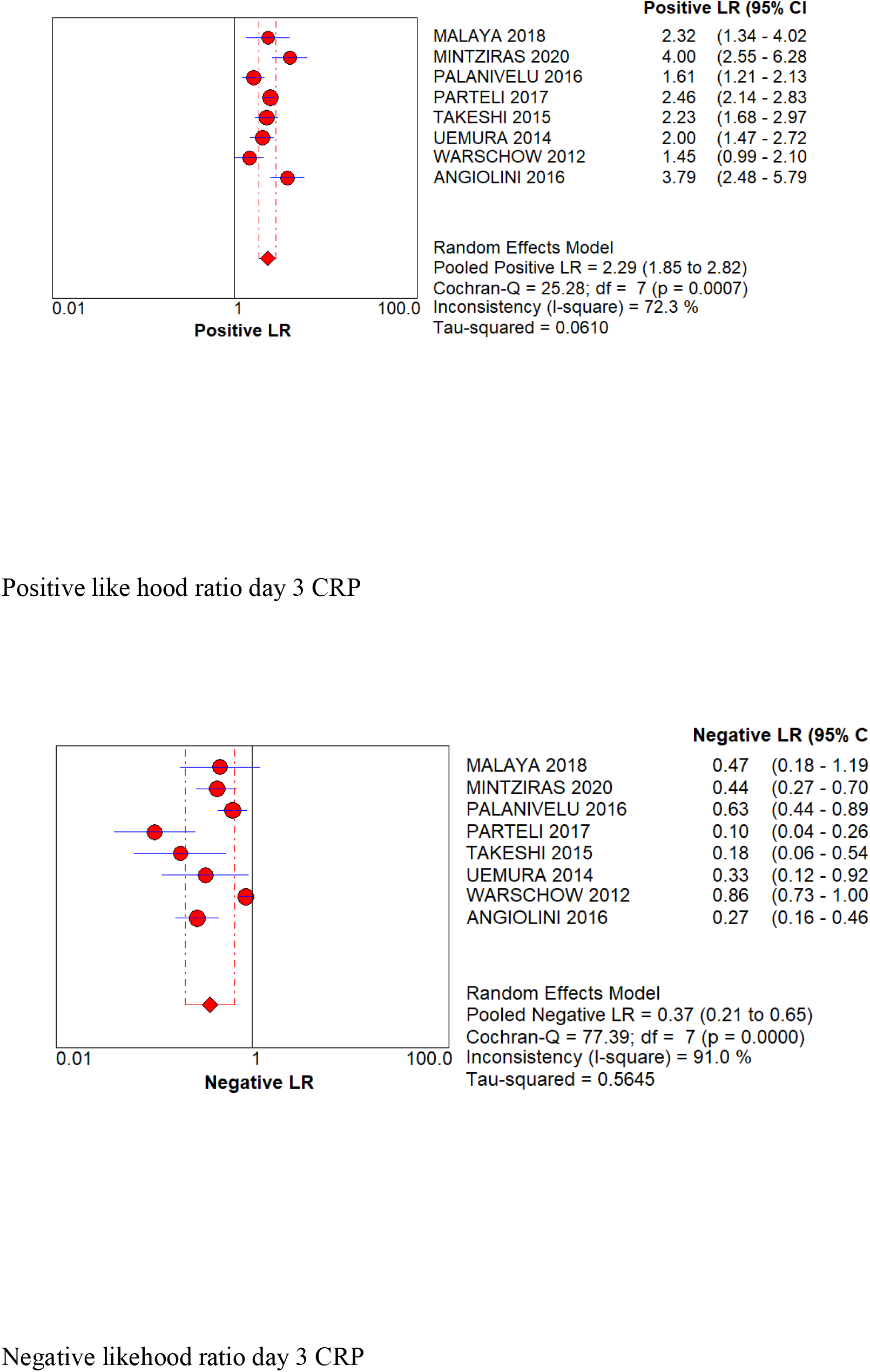
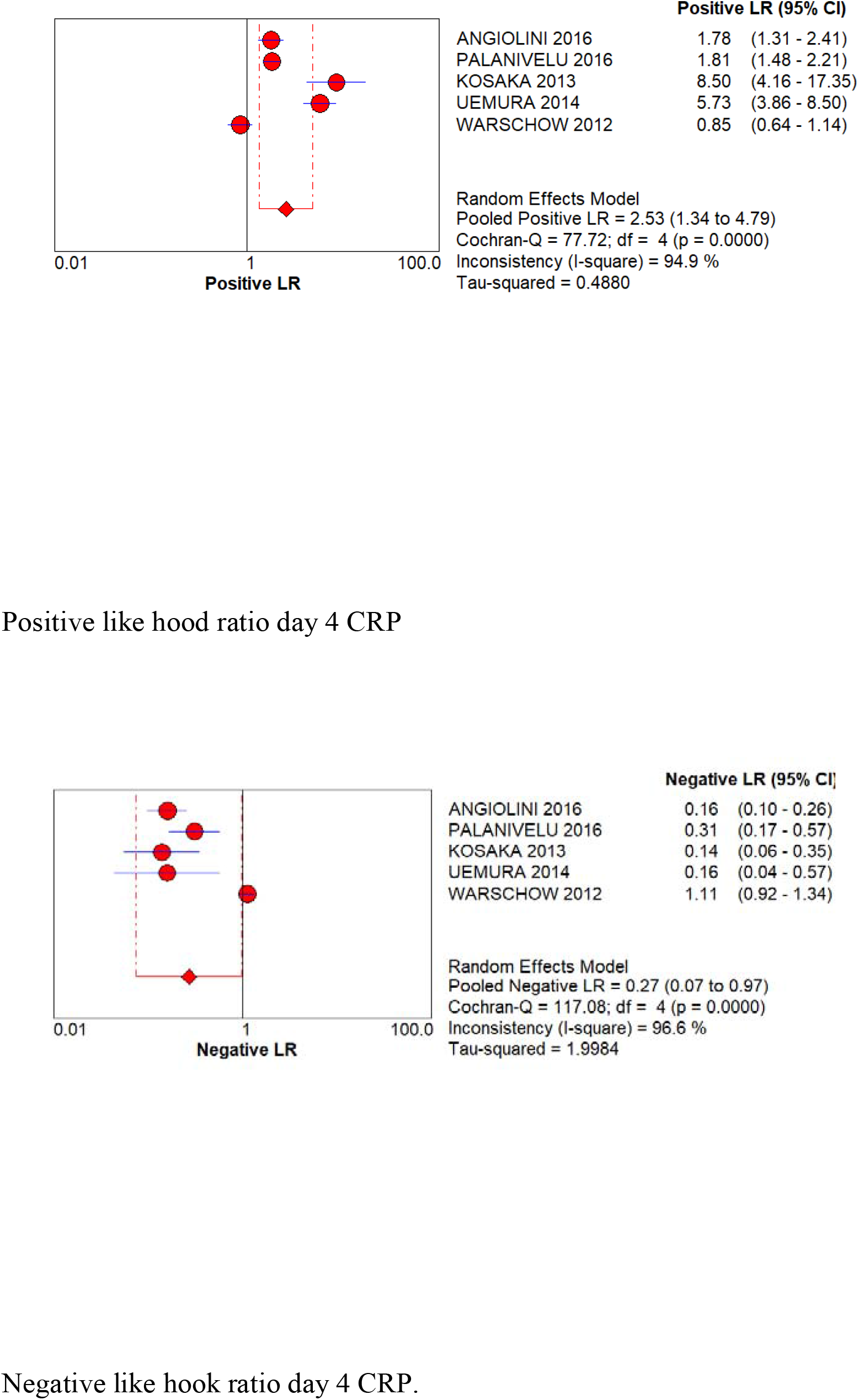
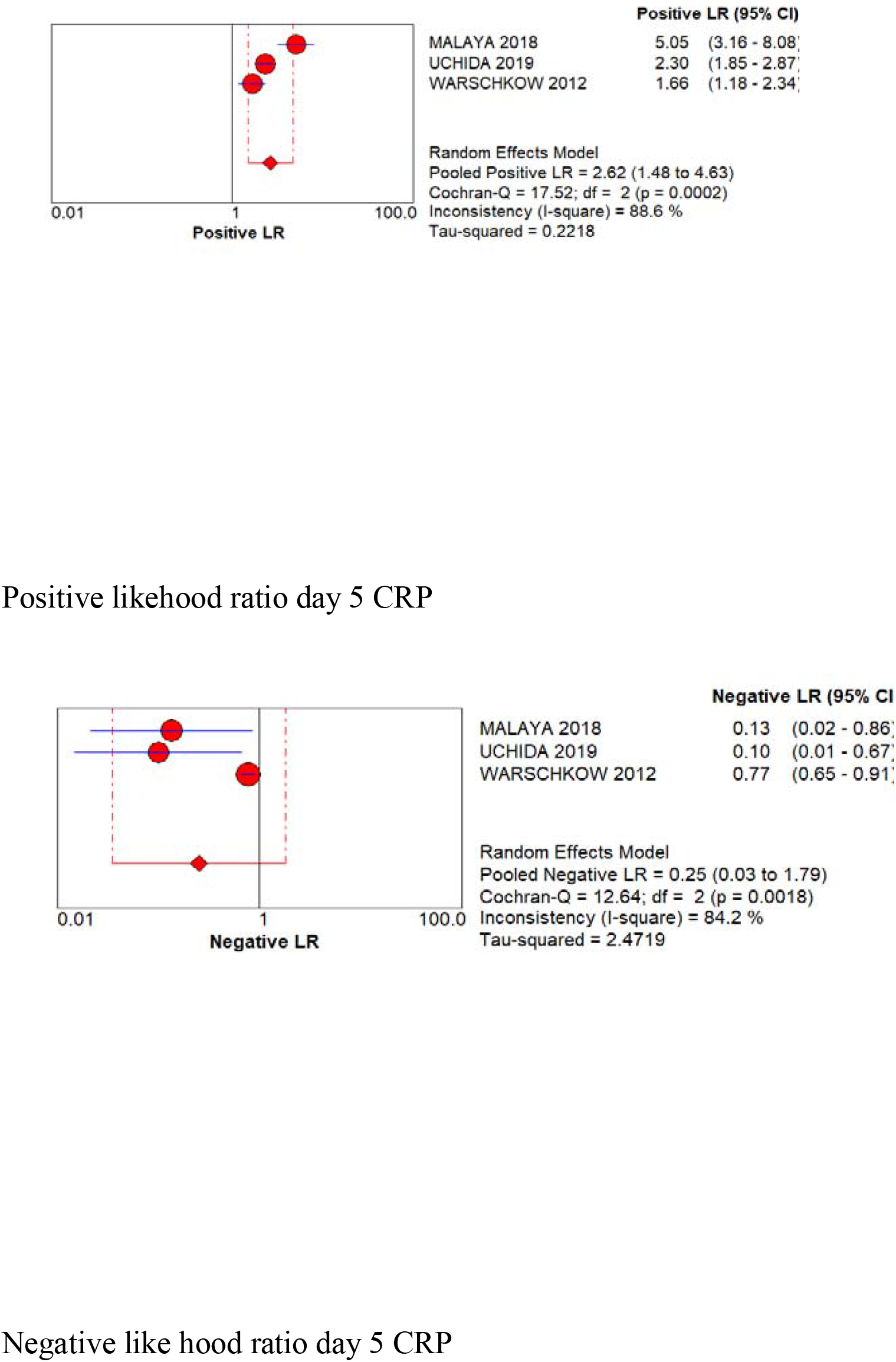
positive and negative like hood ratios of day 3,4,5 CRP.

**Supplement Figure 2.**
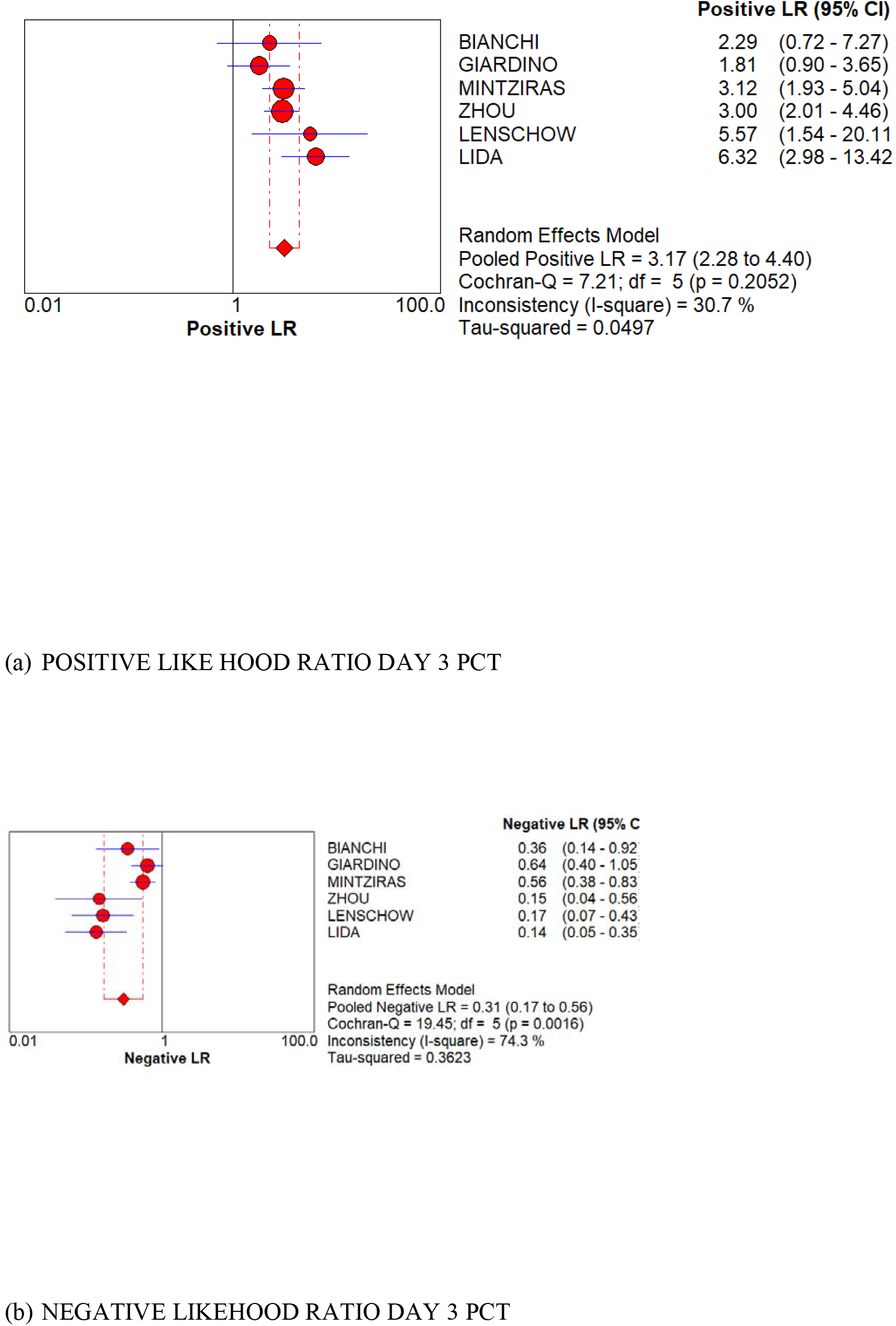
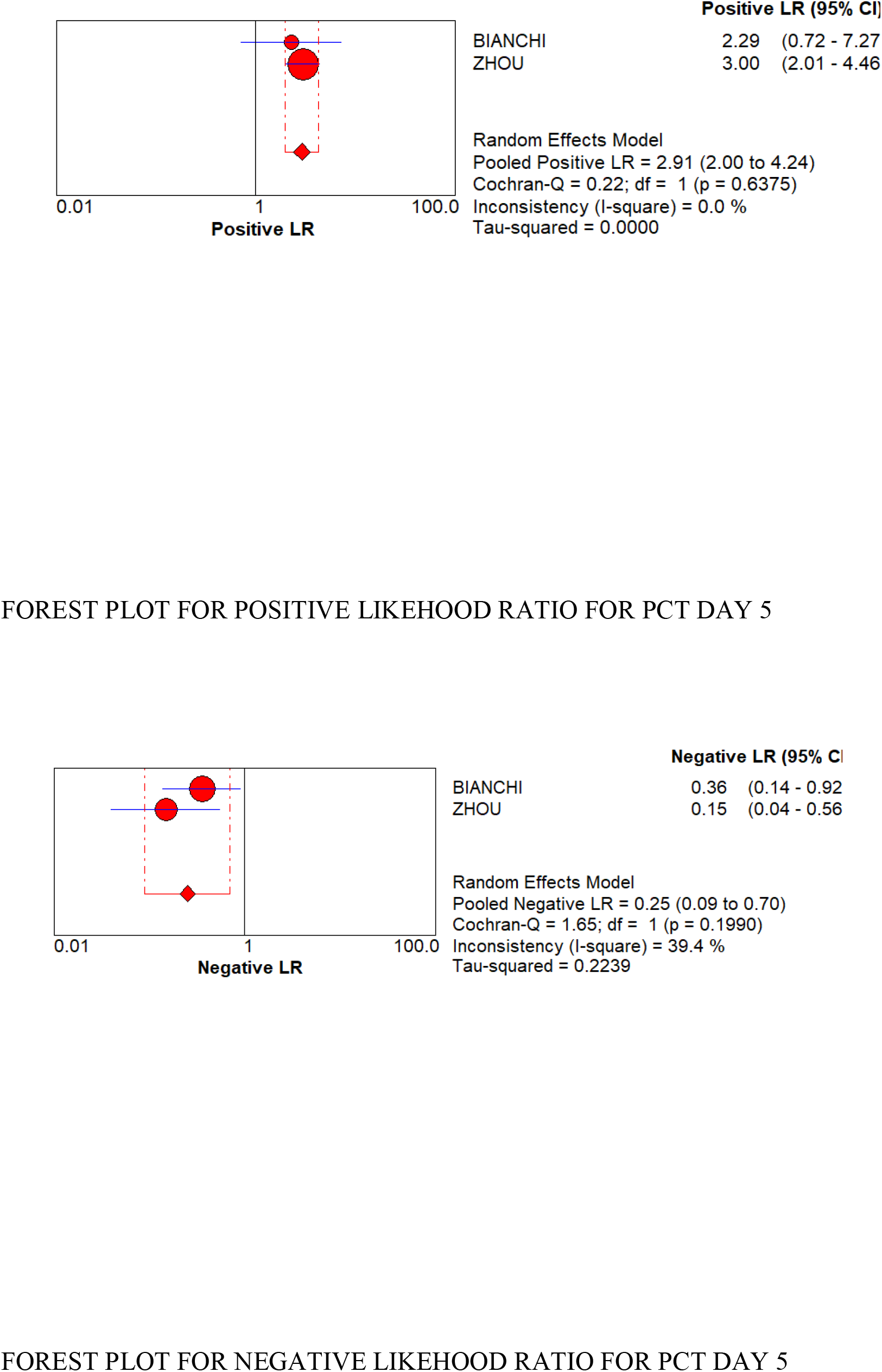
Positive and negative like hood ratio for postoperative day 3 and day 5 CRP and PCT.

## References

1. FuS, ShenS, LiS, HuW, HuaY, KuangM, etal. Riskf actors and outcomes of postoperative pancreatic fistula after pancreatico-duodenectomy: an audit of 532 consecutive cases. BMC Surgery, 2015. 15–34.

2. Krautz C, Nimptsch U, Weber GF, Mansky T, Grützmann R. Effect of Hospital Volume on In-hospital Morbidity and Mortality Following Pancreatic Surgery in Germany. Ann Surg. 2018;267(3):411–417. doi:10.1097/SLA.0000000000002248

3. Barreto SG, Singh A, Perwaiz A, et al. Perioperative antimicrobial therapy in preventing infectious complications following pancreatoduodenectomy. Indian J Med Res. 2017;146(4):514–519. doi:10.4103/ijmr.IJMR_784_15

4. Paniccia A, Hosokawa P, Henderson W, Schulick RD, Edil BH, Mccarter MD, et al. Characteristics of 10-Year Survivors of Pancreatic Ductal Adenocarcinoma. JAMA Surg, 2015. 80045: p1–10.

5. Tan WJ, Ng WQ, Sultana R, et al. Systematic review and meta-analysis of the use of serum procalcitonin levels to predict intra-abdominal infections after colorectal surgery. Int J Colorectal Dis. 2018;33(2):171–180. doi:10.1007/s00384-017-2956-8

6. Spoto S, Valeriani E, Caputo D, et al. The role of procalcitonin in the diagnosis of bacterial infection after major abdominal surgery: Advantage from daily measurement. Medicine (Baltimore). 2018;97(3):e9496. doi:10.1097/MD.0000000000009496

7. Welsch T, Mu□ller SA, Ulrich A, Kischlat A. C-reactive protein as early predictor for infectious postoperative complications in rectal surgery. Surgery, 2007: p1499–507.

8. Facy O, Binquet C, Masson D, Fournel I, Rat P, Ortega-deballon P. Diagnostic Accuracy of Inflammatory Markers As Early Predictors of Infection After Elective Colorectal Surgery-Results From the IMACORS Study. Ann Surg, 2015. p1–6.

9. Lagoutte N, Facy O, Ravoire A, Chalumeau C, Jonval L, Rat P, et al. C-reactive protein and procalcitonin for the early detection of anastomotic leakage after elective colorectal surgery : Pilot study in 100 patients. J. Visc. Surg, 2012. 149: p345–9

10. Warschkow R, Tarantino I, Schmied BM, Steffen T. Diagnostic study and meta-analysis of C-reactive protein as a predictor of post-operative inflammatory complications after gastroesophageal cancer surgery. Langhenbecks Arch Surg, 2012. p727–36.

11. Oberhofer D. Comparison of C-reactive protein and procalcitonin as predictors of postoperative infectious complications after elective colorectal surgery. CMJ, 2012. p612–9.

12. Rowland T, Hilliard H, Barlow G. Procalcitonin: potential role in diagnosis and management of sepsis. Adv Clin Chem. 2015;68:71–86. doi:10.1016/bs.acc.2014.11.005

13. Moher D, Liberati A, Tetzlaff J, Altman DG; PRISMA Group. Preferred reporting items for systematic reviews and meta-analyses: the PRISMA statement. BMJ. 2009 Jul 21;339:b2535. doi: 10.1136/bmj.b2535. PMID: 19622551; PMCID: PMC2714657.

14. Manchikanti L, Datta S, Smith HS, Hirsch JA. Evidence-based medicine, systematic reviews, and guidelines in interventional pain management: part 6. Systematic reviews and meta-analyses of observational studies. Pain Physician. 2009 Sep-Oct;12(5):819–50. PMID: 19787009.

15. Bassi C, Marchegiani G, Dervenis C, Sarr M, Abu Hilal M, Adham M, Allen P, et al. International Study Group on Pancreatic Surgery (ISGPS). The 2016 update of the International Study Group (ISGPS) definition and grading of postoperative pancreatic fistula: 11 Years After. Surgery. 2017 Mar;161(3):584–591. doi: 10.1016/j.surg.2016.11.014. Epub 2016 Dec 28. PMID: 28040257.

16. Whiting PF, Rutjes AW, Westwood ME, Mallett S, Deeks JJ, Reitsma JB, Leeflang MM, Sterne JA, Bossuyt PM, Group Q (2011) QUADAS-2: a revised tool for the quality assessment of diagnostic accuracy studies. Ann Intern Med 155(8):529–536. https://doi.org/10.7326/0003-4819-155-8-201110180-00009

17. van Enst, W.A., Ochodo, E., Scholten, R.J. et al. Investigation of publication bias in meta-analyses of diagnostic test accuracy: a meta-epidemiological study. BMC Med Res Methodol 14, 70 (2014). https://doi.org/10.1186/1471-2288-14-70

18. Freeman MF, John W (1950) Tukey. BTransformations related to the angular and the square root.^ the. Ann Math Stat 21(4):607–611. http://www.jstor.org/stable/2236611. https://doi.org/10.1214/aoms/1177729756

19. Higgins JP, Thompson SG, Deeks JJ, Altman DG (2003) Measuring inconsistency in meta-analyses. BMJ 327(7414):557–560. https://doi.org/10.1136/bmj.327.7414.557

20. Bianchi RA, Haedo AS, Romero MC. Papel de la determinación de procalcitonina plasmática en el seguimiento postoperatorio de la duodenopancreatectomía cefálica. Cir Esp 2006;79:356–360

21. Lenschow C, Hummel R, Lindner K, et al. Procalcitonin--a Marker for Anastomotic Insufficiency after Pancreatoduodenectomy?. Clin Lab. 2016;62(1-2):209–217. doi:10.7754/clin.lab.2015.150703

22. Iida H, Maehira H, Mori H, Tani M. Serum procalcitonin as a predictor of infectious complications after pancreaticoduodenectomy: review of the literature and our experience. Surg Today. 2020;50(2):87–96. doi:10.1007/s00595-019-01811-y

23. Giardino A, Spolverato G, Regi P, et al. C-Reactive Protein and Procalcitonin as Predictors of Postoperative Inflammatory Complications After Pancreatic Surgery. J Gastrointest Surg. 2016;20(8):1482–1492. doi:10.1007/s11605-016-3171-6

24. Zhou Q, Xia Y, Lei Z. The predictive value of procalcitonin for postoperative early pancreatic fistula. BMC Surg. 2020;20(1):90. Published 2020 May 6. doi:10.1186/s12893-020-00755-2.

25. Mintziras I, Maurer E, Kanngiesser V, Bartsch DK. C-reactive protein and drain amylase accurately predict clinically relevant pancreatic fistula after partial pancreaticoduodenectomy. Int J Surg. 2020;76:53–58. doi:10.1016/j.ijsu.2020.02.025.

26. Malya FU, Hasbahceci M, Tasci Y, Kadioglu H, Guzel M, Karatepe O, et al. The Role of C-Reactive Protein in the Early Prediction of Serious Pancreatic Fistula Development after Pancreaticoduodenectomy. Gastroenterol Res Pract. 2018 Jan 28;2018:9157806. doi: 10.1155/2018/9157806. PMID: 29619047; PMCID: PMC5830281.

27. Partelli S, Pecorelli N, Muffatti F, Belfiori G, Crippa S, Piazzai F, et al. Early Postoperative Prediction of Clinically Relevant Pancreatic Fistula after Pancreaticoduodenectomy: usefulness of C-reactive Protein. HPB (Oxford). 2017 Jul;19(7):580–586. doi: 10.1016/j.hpb.2017.03.001. Epub 2017 Apr 7. PMID: 28392159.

28. Angiolini MR, Gavazzi F, Ridolfi C, Moro M, Morelli P, Montorsi M, et al. Role of C-Reactive Protein Assessment as Early Predictor of Surgical Site Infections Development after Pancreaticoduodenectomy. Dig Surg. 2016;33(4):267–75. doi: 10.1159/000445006

29. Uemura K, Murakami Y, Sudo T, Hashimoto Y, Kondo N, Nakagawa N, et al. Indicators for proper management of surgical drains following pancreaticoduodenectomy. J Surg Oncol. 2014 Jun;109(7):702–7. doi: 10.1002/jso.23561.

30. Palani Velu LK, McKay CJ, Carter CR, McMillan DC, Jamieson NB, Dickson EJ. Serum amylase and C-reactive protein in risk stratification of pancreas-specific complications after pancreaticoduodenectomy. Br J Surg. 2016 Apr;103(5):553–63. doi: 10.1002/bjs.10098

31. Kosaka H, Kuroda N, Suzumura K, Asano Y, Okada T, Fujimoto J. Multivariate logistic regression analysis for prediction of clinically relevant pancreatic fistula in the early phase after pancreaticoduodenectomy. J Hepatobiliary Pancreat Sci. 2014 Feb;21(2):128–33. doi: 10.1002/jhbp.11.

32. Uchida Y, Masui T, Nakano K, Yogo A, Yoh T, Nagai K, et al. Combination of postoperative C-reactive protein value and computed tomography imaging can predict severe pancreatic fistula after pancreatoduodenectomy. HPB (Oxford). 2020 Feb;22(2):282–288. doi: 10.1016/j.hpb.2019.06.020.

33. Takeishi K, Maeda T, Yamashita Y, Tsujita E, Itoh S, Harimoto N, Ikegami T, Yoshizumi T, Shirabe K, Maehara Y. A Cohort Study for Derivation and Validation of Early Detection of Pancreatic Fistula After Pancreaticoduodenectomy. J Gastrointest Surg. 2016 Feb;20(2):385–91. doi: 10.1007/s11605-015-3030-x.

34. Cousin F, Ortega-Deballon P, Bourredjem A, Doussot A, Giaccaglia V, Fournel I. Diagnostic Accuracy of Procalcitonin and C-reactive Protein for the Early Diagnosis of Intra-abdominal Infection After Elective Colorectal Surgery: A Meta-analysis. Ann Surg. 2016;264(2):252–256.doi:10.1097/SLA.0000000000001545.

35. Rhodes, Andrew MB BS, MD(Res) (Co-chair)^1^; Evans, Laura E. MD, MSc, FCCM (Co-chair)^2^; Alhazzani, Waleed MD, MSc, FRCPC (methodology chair)^3^; Levy, Mitchell M. MD, MCCM^4^; Antonelli, Massimo MD^5^; Ferrer, Ricard MD, PhD^6^ et al. Management of Sepsis and Septic Shock: 2016, Critical Care Medicine: March 2017 - Volume 45 - Issue 3 - p 486–552 doi: 10.1097/CCM.0000000000002255.

36. Giaccaglia V, Salvi PF, Antonelli MS, Nigri G, Pirozzi F,Casagranda B, Giacca M, Corcione F, de Manzini N, Balducci G, Ramacciato G (2016) Procalcitonin reveals early dehiscence incolorectal surgery: the PREDICS study. Ann Surg 263(5):967–972. https://doi.org/10.1097/SLA.0000000000001365.

37. Garcia-Granero A, Frasson M, Flor-Lorente B, Blanco F, Puga R,Carratala A, Garcia-Granero E (2013) Procalcitonin and C reactive protein as early predictors of anastomotic leak in colorectal surgery:a prospective observational study. Dis Colon Rectum 56(4):475–483. https://doi.org/10.1097/DCR.0b013e31826ce825.

38. Giaccaglia V, Salvi PF, Cunsolo GV, Sparagna A, Antonelli MS, Nigri G, Balducci G, Ziparo V (2014) Procalcitonin, as an early biomarker of colorectal anastomotic leak, facilitates enhanced recovery after surgery. J Crit Care 29(4):528–532. https://doi.org/10.1016/j.jcrc.2014.03.036.

